# A systematic analysis of splicing variants identifies new diagnoses in the 100,000 Genomes Project

**DOI:** 10.1101/2022.01.28.22270002

**Authors:** Alexander J.M. Blakes, Htoo Wai, Ian Davies, Hassan E. Moledian, April Ruiz, Tessy Thomas, David Bunyan, N Simon Thomas, Christine P. Burren, Lynn Greenhalgh, Melissa Lees, Amanda Pichini, Sarah F. Smithson, Ana Lisa Taylor Tavares, Peter O’Donovan, Andrew G.L. Douglas, Genomics England Research Consortium, Splicing and Disease Working Group, Nicola Whiffin, Diana Baralle, Jenny Lord

## Abstract

Genomic variants which disrupt splicing are a major cause of rare genetic disease. However, variants which lie outside of the canonical splice sites are difficult to interpret clinically. Here, we examine the landscape of splicing variants in whole-genome sequencing data from 38,688 individuals in the 100,000 Genomes Project, and assess the contribution of non-canonical splicing variants to rare genetic diseases. We show that splicing branchpoints are highly constrained by purifying selection, and harbour damaging non-coding variants which are amenable to systematic analysis in sequencing data. From 258 *de novo* splicing variants in known rare disease genes, we identify 35 new likely diagnoses in probands with an unsolved rare disease. We use phenotype matching and RNA studies to confirm a new diagnosis for six individuals to date. In summary, we demonstrate the clinical value of examining non-canonical splicing variants in participants with unsolved rare diseases.

## Introduction

Improved diagnosis of rare genetic diseases remains a significant clinical and research challenge^1^. Diagnostic yields in individuals with rare diseases remain below 50%, despite extensive investigations including whole-genome sequencing^2^. The accurate interpretation of genomic variants in existing sequencing data presents an important opportunity to narrow the diagnostic gap^3^.

Splicing is the process by which introns are removed from a pre-mRNA primary transcript. Almost all human protein coding genes are spliced, and disruption of splicing is a major cause of rare genetic diseases^4^. The improved interpretation of splicing variants is therefore a major opportunity to improve clinical outcomes for individuals with undiagnosed rare disease^5^.

Already, “canonical splice site “ (CSS) variants within 2bp of an exon-intron junction are widely annotated as “loss of function “ (LoF) variants, and are known to be strong diagnostic candidates in “loss-of function “ disorders^6^. The contribution of non-canonical splicing variants to rare disease is also becoming increasingly recognised^7^. Up to 27% of pathogenic *de novo* splicing variants in exome-sequencing data are found in non-canonical positions^8^. Several studies^7–9^ have developed the concept of a “near-splice “ region, usually tens of base pairs around an exon-intron junction, which contains many conserved splicing motifs.

However, near-splice variants are under-reported in clinical databases^8^, and no standards exist for their interpretation. Furthermore, variants distal to the near-splice region, including branchpoint variants and deep intronic variants, can also disrupt splicing, and their overall contribution to rare disease is unknown. Individual instances of pathogenic branchpoint variants have been previously described^10,11^, but they have not been systematically characterised in a large rare disease cohort.

Recently, large population genomic datasets have provided the statistical power necessary to measure constraints on genetic variation within human populations. One powerful metric which uses this approach is the mutability-adjusted proportion of singletons (MAPS)^12^, which identifies classes of variation which are subject to purifying selection, and are therefore likely to be deleterious. MAPS has previously been calculated in many contexts, including for near-splice positions in the Exome Aggregation Consortium (ExAC)^8^, and for upstream start-codon-creating variants in Genome Aggregation Database (gnomAD)^13^.

Recent advances in computation and artificial intelligence have led to the development of numerous *in silico* predictors for the prioritization of splicing variants^14^. For example, SpliceAI is a machine learning tool which robustly predicts splice sites and splice-disrupting variants^15^, and out-performs other algorithms in predicting splicing consequences from sequence data^16^. However, in clinical variant interpretation, well-validated functional assays have greater weight than *in silico* predictions of variant effect^6^, and functional validation of most splicing variants is still required to confirm a molecular diagnosis.

Here, we perform a systematic analysis of splicing variants in whole-genome sequencing data from 38,688 individuals in the Rare Disease arm of the 100,000 Genomes Project (100KGP)^17^. We evaluate the contribution of canonical, near-splice, and splicing branchpoint variants to rare genetic diseases in this cohort. We show that splicing branchpoints harbour deleterious non-coding variants which are amenable to systematic analysis in WGS data. We used a gene-agnostic approach to prioritise 258 *de novo* splicing variants in families affected by a rare genetic disorder. Of these, at least 84 were already considered to be diagnostic, and we identified an additional 35 variants which are likely to be diagnostic given the available molecular, phenotypic, and *in silico* data. We confirmed a new molecular diagnosis for six participants, including four out of five participants for whom RNA studies were performed. Ultimately, we demonstrate the clinical and diagnostic value of examining both canonical and non-canonical splicing variants in unsolved rare diseases.

## Subjects and Methods

### Cohort, sequencing, and tiering

This analysis was performed on whole-genome sequencing data from 38,688 participants in the Rare Disease arm of the 100,000 Genomes Project^18^. These comprised 26,660 unaffected parents of rare disease probands, and 12,028 participants (offspring) for whom trio WGS data was available. Only participants for whom WGS data was aligned to GRCh38 were included in this study. Parents affected by a rare genetic disease were excluded from the analysis of variant constraint (see below). Otherwise, participants were not selected or stratified by any other criterion. The sequencing and bioinformatic pipelines, as well as the “tiering “ framework for variant prioritisation, have been previously described^17^. Briefly, variants meeting filtering criteria and falling within applied virtual gene panels were annotated as tier 1 (loss of function or *de novo* protein-altering variants), tier 2 (other variant types eg missense, with correct mode of inheritance), or tier 3 (all other filtered variants). For example, CSS variants in an appropriate gene panel are annotated as tier 1.

### Defining CDS exons and near-splice positions

We identified coding sequences in high-confidence protein-coding transcripts from GENCODE v29^19^ (GRCh38) using the following filtering criteria: feature type = “CDS “, gene_type = “protein_coding “, transcript_type = “protein_coding “, level != “level 3 “, and tag = “CCDS “, “appris_principal_1 “, “appris_candidate_longest “, “appris_candidate “, or “exp_conf “. 401,314 CDS features (207,548 unique) met these criteria. Only autosomal CDS were included in the subsequent analyses. UTR and other non-coding exons were not included in this analysis.

For each CDS feature, we annotated individual genomic positions with their positions relative to a splice donor or acceptor site, excluding any sites with conflicting annotations. We defined the near-splice region around the acceptor site as 25bp of intronic sequence (acceptor -25 to acceptor -1), and 11bp of exonic sequence (acceptor 0 to acceptor +10). Around the donor site, we included 11bp of exonic sequence (donor -10 to donor 0) and 10 bp of intronic sequence (donor +1 to donor +10). 9,588,491 distinct near-splice positions were identified.

### phyloP

We annotated each near-splice position with phyloP scores from multiple alignments of 99 vertebrate species to the human genome (phyloP 100-way)^20^ with pyBigWig, an open-source Python package^21^, using BigWig files downloaded from the UCSC genome browser (hg38)^22,23^.

### SpliceAI

For every possible near-splice SNV in our positions of interest (i.e. all three possible single base changes at each of the 9,588,491 positions), we annotated the predicted effect on splicing with SpliceAI^15^. We annotated variants with pre-computed genome-wide SpliceAI v1.3 scores (available via https://github.com/Illumina/SpliceAI) using BCFtools v1.9^24^. A SpliceAI annotation was available for 28,265,193 variants (98.2% of 28,765,473 possible variants).

Aggregate SpliceAI scores for each near-splice position were calculated as the mean probability that any variant at this position disrupts splicing. The probability that a given variant disrupts splicing was calculated as the probability (P) of any one of the SpliceAI-predicted splicing events occurring, (i.e. 1 - probability of *no* events occurring). The predicted splicing events are acceptor gain (AG), acceptor loss (AL), donor gain (DG), or donor loss (DL), giving:

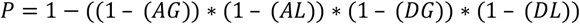

### Mutability-adjusted proportion of singletons

In addition to the near-splice SNVs identified above, we also determined the set of all possible coding SNVs in our exons of interest. These were annotated with the reference base for each position (GRCh38, GenBank assembly accession GCA_000001405.15) and its immediate sequence context (1bp either side) with bedtools version 2.27.1^25^.

We annotated every possible coding SNV within our exons of interest with the Variant Effect Predictor (VEP) version 99^26^. For each variant, only the consequence in one transcript (typically the canonical transcript, as determined by VEP ‘s “–pick “ flag), was used. Only synonymous, missense, and nonsense variants were included in the subsequent analysis. Synonymous variants within a near-splice region were classed as near-splice variants for the MAPS calculation and were excluded from the synonymous variant set. Missense variants within a near-splice region were excluded from the analysis altogether. Nonsense variants within a near-splice region were classed as nonsense variants and excluded from the near-splice variant set.

We interrogated whole genome sequencing data from 26,660 unaffected parents in the Genomics England (GEL) Rare Disease cohort for SNVs overlapping the near-splice and coding positions defined above using BCFtools v1.9^24^. Only variants passing all filters (see https://research-help.genomicsengland.co.uk/display/GERE/aggV2+Details) within the GEL aggregated multi-sample VCF were included. We identified 915,024 synonymous, 1,965,441 missense, 53,825 nonsense, and 672,528 near-splice variants, and calculated allele counts across the 26,660 unaffected parents for each variant.

We calculated MAPS with custom Python scripts, adapting code written in R by Patrick J. Short^27^ (https://github.com/pjshort/dddMAPS). We used the mutation rate of a given trinucleotide context calculated by Samocha et al^28^. The proportion of singletons for each position was adjusted for the mutability of the immediate sequence context using a linear model trained on synonymous variants within the same exons.

### Branchpoints

Splicing branchpoint positions were identified by LaBranchoR, a machine-learning tool trained on experimentally-validated branchpoints, which accurately identifies at least one branchpoint for the majority of introns genome-wide^29^. Pre-computed LaBranchoR scores are publicly available for every position 1-70bp upstream of a splice acceptor (GENCODE v19, hg19)^29^. For each intron, the highest scoring position was annotated as the branchpoint (BP), totalling 195,863 putative branchpoints.

We converted each branchpoint to hg38 coordinates using the UCSC Liftover tool^30^. We annotated five positions upstream (−5 to -1) and five positions downstream (+1 to +5) of each branchpoint, as well as every possible SNV at each of these positions using custom Python scripts. phyloP scores for each position, and SpliceAI scores for each variant, were determined as above. We calculated the MAPS statistic for these branchpoint positions in the same cohort of participants as described above. Comparison of MAPS scores in branchpoint positions was made to the same set of coding variants as described above.

### Statistics

The null hypotheses that near-splice and branchpoint MAPS scores did not significantly differ from synonymous variants were tested with two-sided Chi squared tests of the observed vs the expected number of singletons in each variant class. In order that the synonymous MAPS did not equal zero, all MAPS scores were first corrected by addition of the synonymous unadjusted proportion of singletons. For each variant class the “observed “ proportion of singletons was taken as the number of alleles multiplied by the corrected MAPS score for that variant class. The “expected “ number of singletons was taken as the number of alleles multiplied by the corrected MAPS score for synonymous variants. Multiple testing was accounted for by Bonferroni correction: 79 tests at alpha = 0.05 gave a significance threshold of <6.3×10^−4^.

### *De novo* variants

*De novo* mutations (DNMs) overlapping near-splice positions were identified from a set of 1,004,599 high confidence *de novo* calls in 13,949 trios from 12,609 rare disease families. The annotation pipeline used to identify these variants is publicly available^31^. Briefly, a multisample VCF for each trio was annotated for putative DNMs using custom scripts. Putative DNMs were then filtered by a series of “Global “, “Base “, and “Stringent “ filters (see reference^31^). Unless otherwise stated, our analyses were performed on DNMs aligned to GRCh38 (870,559 DNMs in 12,028 trios).

At the outset of this project this dataset was not available. Preliminary work to identify candidate diagnostic *de novo* variants was undertaken in a smaller set of 402,464 variants identified through a custom filtering strategy by Patrick J. Short (Wellcome Sanger Institute, personal correspondence). These variants were identified by applying post-processing filters to DNMs in 4,967 trios identified by the GEL Platypus variant caller and aligned to GRCh38. They were filtered according to the following criteria: genotype heterozygous in offspring and homozygous reference in both parents, no more than one alternate allele read in either parent, variant allele frequency in the offspring between 0.3 and 0.7, greater than 20 sequencing reads in the offspring and both parents, fewer than 98 sequencing reads in the offspring, no overlap with locus control regions, no overlap with hg38 “patch regions “, no other DNM within 20bp in the same individual. Some candidate variants identified in this preliminary dataset are not present in the larger *de novo* set, owing to differences in the filtering pipeline. Unless explicitly stated, the data presented here are from the larger DNM set, above.

### Candidate diagnostic variants

To identify candidate diagnostic near-splice and branchpoint variants, we annotated all GRCh38 autosomal *de novo* SNVs passing the “stringent “ filters (above) with VEP (version 99). For each variant, only the consequence in one transcript per gene (determined by VEP ‘s “--per_gene “ flag) was considered. We annotated these variants with SpliceAI as described above. We filtered for variants overlapping our branchpoint or near-splice positions of interest, finding 3,672 such variants. Where a variant had both a branchpoint and a near-splice annotation, only the near-splice annotation was kept. We then filtered for variants overlapping any known monoallelic rare disease gene with a loss of function mechanism using the G2P DD, G2P Eye, and G2P Skin gene lists^32^ (accessed 27/10/2021, confirmed and probable genes only). In total we identified 258 candidate splicing DNMs (238 near-splice, 20 branchpoint) in 255 participants, across 137 genes.

To identify new diagnoses in the cohort, we annotated these variants with tiering data, phenotype data, and participant outcome data from the GEL bioinformatics pipeline^33^. For each participant and DNM, we manually reviewed the similarity between the HPO terms recorded at recruitment and the phenotype expected for a loss-of-function variant in that gene. Excluding any participants whose case was already solved through 100KGP, we identified 35 new likely diagnostic variants with at least a plausible phenotype match. In each instance, we placed a clinical collaboration request with Genomics England to recruit the participant to the Splicing and Disease Study for functional characterisation of the variant.

Genomics England does not allow re-identification of participants outside of a secure research environment. In order to protect participant identities, the HPO terms given here are “abstracted “ by moving up one level in the HPO hierarchy. For example “Tetralogy of Fallot “ becomes “Conotruncal defect “.

### Functional validation

Samples from five participants underwent functional characterisation through the Splicing and Disease study at The University of Southampton. Blood was collected in PAXgene Blood RNA tubes, with the PAXgene Blood RNA kit (PreAnalytix, Switzerland) used to extract RNA. Random hexamer primers were used to synthesise complementary DNA (cDNA) by reverse transcription using the High-Capacity cDNA Reverse Transcription kit (ThermoFisher Scientific).

Reverse transcription polymerase chain reaction (RT-PCR) was used to test for splicing alterations. Primers were designed for each variant to include at least two exons up- and downstream of the target (primer sequences available upon request). Agarose gel electrophoresis was used to assess participant vs control PCR products, and purified PCR products were analysed by Sanger sequencing.

### Ethics

The 100,000 Genomes Project Protocol has ethical approval from the HRA Committee East of England – Cambridge South (REC Ref 14/EE/1112). This study was registered with Genomics England underResearch Registry Projects 143, 165, and 166. The Splicing and Disease study has ethical approval from the Health Research Authority (IRAS Project ID 49685, REC 11/SC/0269) and The University of Southampton (ERGO ID 23056), with informed consent given for splicing studies in a research context.

### Code

All analyses were performed within a protected research environment which is available to registered users (see https://www.genomicsengland.co.uk/about-gecip/for-gecip-members/data-and-data-access/). Clinical data and tiering data were accessed through the LabKey application within this research environment. The analyses were performed using Python 3.7.6 and Pandas 1.2.1. Figures were generated in R 3.5.2 using RStudio 1.1.463. All code is available online at https://github.com/alexblakes/100KGP_splicing.

## Results

### Signals of constraint at near-splice positions are replicated in a large healthy cohort

To estimate the deleteriousness of variation in near-splice positions, we calculated aggregate measures of evolutionary conservation, selective constraint, and pathogenicity prediction for nucleotides within near-splice regions genome-wide.

Evolutionary conservation was measured by base-wise phyloP score. The CSSs are very highly conserved (mean phyloP = 6.34) (Figure 1). Other intronic splicing positions with high phyloP scores include the D+5 (3.44), D+4 (2.39), D+3 (1.97), A-3 (1.70), and D+6 (1.29) sites. Notably, the A-4 position is very weakly conserved (0.076). Coding positions are generally more highly conserved than intronic sequences. The redundancy of third codon positions and bias for in-phase exons^34^ is reflected in lower phyloP scores at every third position, except for the donor 0 position (mean phyloP = 5.01), which is more highly conserved than any other coding position.

**Figure 1.**
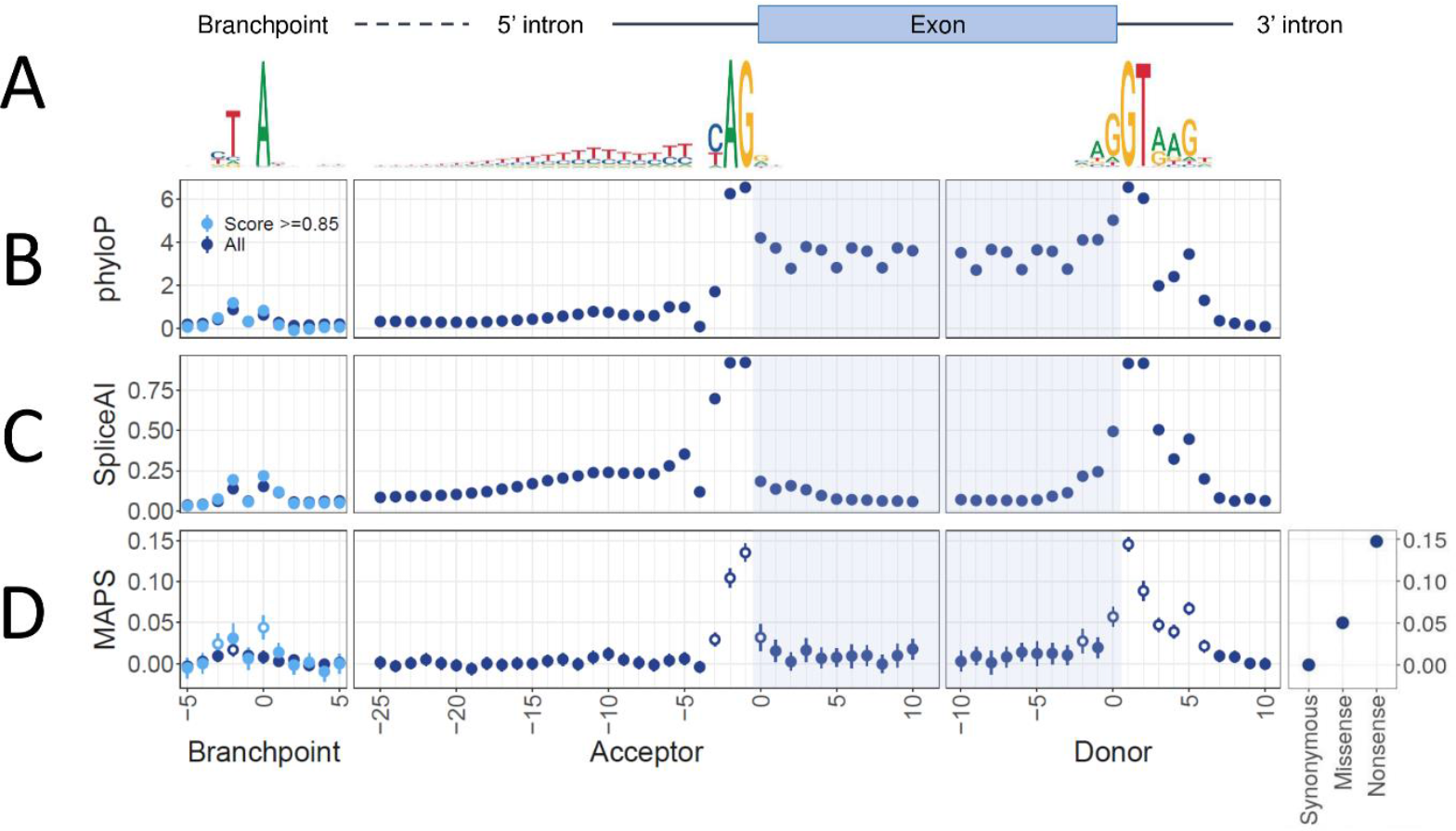
Conservation, predicted splice disruption, and constraint at near-splice and branchpoint positions across 207,548 CDS features in protein coding genes. **A:** Position-weight matrices and schematic indicating position of conserved splicing motifs relative to exon / intron boundaries. **B:** Mean phyloP 100-way scores at splicing positions. Error bars indicate 95% confidence intervals. **C:** SpliceAI scores for all possible near splice SNVs. Scores represent the mean probability that any variant at this position disrupts splicing, as predicted by SpliceAI (see Methods). Error bars represent the 95% confidence interval. **D:** Mutability-adjusted proportion of singletons (MAPS) for both coding and near-splice SNVs. Error bars indicate 95% confidence intervals. Positions with a significantly higher MAPS than synonymous variants are indicated with open circles (see Methods). For branchpoint positions, dark blue points represent all putative branchpoints, whereas light blue points represent branchpoints with a LaBranchoR score >0.85

To measure selective constraint at near-splice positions, we calculated the degree of purifying selection acting at near-splice positions using MAPS^12^. MAPS was calculated across near splice positions genome-wide, using every observed synonymous, missense, nonsense, and near-splice SNV in 207,548 distinct CDS exons for 26,660 unaffected parents in the 100KGP Rare Disease cohort. The most significant signals of purifying selection are at the CSS, with MAPS scores of 0.089–0.146 (p<1.2×10^−43^), approaching those of nonsense variants (0.16) (Figure 1). The non-canonical positions with a MAPS score significantly above the synonymous baseline after Bonferroni correction include the D-2, D0, D+3, D+4, D+5, D+6, A-3 and A+1 positions (p<6.3×10^−4^). The MAPS scores at D0 and D+5 variants (MAPS = 0.057 and 0.067 respectively) are comparable to that at missense variants (0.052). These results are highly concordant with previous near-splice MAPS calculations in the DDD and ExAC datasets^8^.

### A subset of splicing branchpoints are highly constrained

Having replicated earlier findings^8^ in our cohort, we expanded our analysis to examine splicing branchpoints, which have not been previously characterised using MAPS.

We repeated our analysis of conservation, constraint, and SpliceAI predicted pathogenicity using a set of 195,863 putative branchpoints predicted by LaBranchoR^29^, a deep-learning tool trained on experimentally-validated branchpoints.

Annotating each position with base-wise phyloP scores, we found modest conservation of BP0 (0.62) and BP-2 (0.87), consistent with previous results^29^ (Figure 1).

Next, we calculated the MAPS statistic for these branchpoint positions in the same cohort described above. When all putative branchpoints were considered, only the BP-2 position has a significantly higher MAPS score than the synonymous baseline (MAPS = 0.017, p=1.3 × 10^−4^) (Figure 1). However, when only the most confident branchpoints are considered (LaBranchoR score >0.85, n=57,342), the BP0 (MAPS = 0.044, p=7.6×10^−9^) and BP-3 (0.024, p = 3.7 × 10^−4^) are also significantly constrained. These data suggest that LaBranchoR-predicted branchpoints are functionally important, and that variants near branchpoints may be a significant cause of rare disease.

Next, we calculated SpliceAI scores for every possible SNV around each branchpoint. Again, variants at BP0 (0.15) and BP-2 (0.14) are nominally more likely to disrupt splicing than synonymous coding variants (Figure 1). This trend is more pronounced when only the most confident branchpoints (LaBranchoR score >0.85, n = 57,342) are considered (mean SpliceAI BP0 = 0.22, BP-2 = 0.19).

### New diagnostic candidates among near-splice *de novo* mutations

Having described three orthogonal metrics which independently suggest that certain near-splice and branchpoint variants may be deleterious, we sought to identify new candidate diagnostic variants at these positions.

We interrogated a set of 870,559 DNMs in 12,028 trios for potentially diagnostic splicing variants. We identified 258 *de novo* SNVs overlapping near-splice or branchpoint regions of known monoallelic “loss of function “ rare disease genes in 255 individuals (Supplementary Table 1). Of these, 238 were in near-splice positions, and 20 were within 5bp of a putative branchpoint. (12 variants had both a splice acceptor and a branchpoint annotation; in these cases only the splice acceptor annotation (e.g. A-25) was kept).

Reviewing tiering data from the 100KGP bioinformatics pipeline, we found that of these 258 variants, 83 (32%) were “Tier 1 “, 46 (18%) were “Tier 3 “, and 129 (50%) were not tiered (Supplementary Figure1). Of 59 CSS variants, 36 (61%) were “Tier 1 “, nine (15%) were “Tier 3 “, and 14 (24%) were not tiered. Of ten donor +5 variants, four were “Tier 1 “, two were “Tier 3 “ and four were not tiered (Supplementary Figure 1). Annotation of these variants with SpliceAI generally highlighted variants at positions with high MAPS scores (Supplementary Figure 2).

212 participants with a near-splice DNM had outcome data in the form of “exit questionnaires “ from their referring Genomic Medicine Centre. In 84/111 (76%) of solved cases, the diagnostic variant matched our near-splice finding (Figure 2). This result gives us confidence in our approach to candidate variant identification. Nevertheless, a significant proportion of participants with completed exit questionnaires had unsolved cases (101/212, 48%). These included nine with a DNM in the CSS of a known rare disease gene, one with a donor 0 variant, and four with donor +5 variants, which have previously been estimated to have a 90% positive predictive value in rare disease diagnosis^8^ (Figure 2).

**Figure 2.**
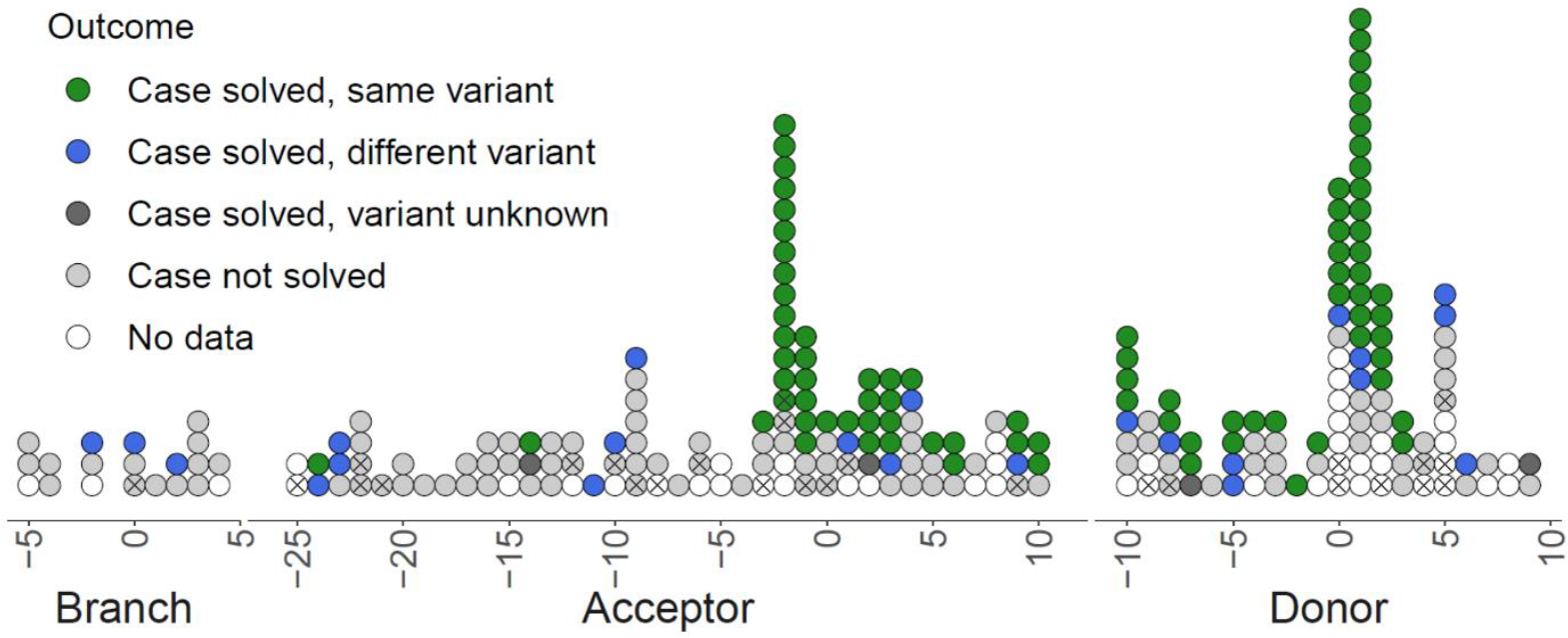
Participant outcomes for rare disease probands with *de novo* splicing variants in known monoallelic loss-of-function rare disease genes. Each point represents a DNM in a rare disease proband. Points are coloured by the clinical outcome for that individual. Crosses indicate variants which were identified as likely new diagnoses in this study. Where a variant overlaps both a branchpoint and a splice acceptor position, only the splice acceptor annotation is given.

For each participant and DNM, we manually reviewed the similarity between the HPO terms recorded at recruitment and the phenotype expected for a loss-of-function variant in that gene. Excluding any participants whose case was already solved through 100KGP, we identified 35 new likely diagnostic variants with at least a plausible phenotype match (Supplementary Table 2). In each instance, we placed a clinical collaboration request with Genomics England to recruit the participant to the Splicing and Disease Study for functional characterisation of the variant.

### New diagnoses among the cohort

Whole blood RNA samples were obtained for five participants with near splice DNMs. RT-PCR was used to characterise the splicing impact of each variant (Supplementary Figure 3). Abnormal splicing events (all exon skipping) were detected in four participants (participants 74 (*ARID1A*, A-3), 249 (*USP7*, D+5), 259 (*TLK2*, D+5), 261 (*KAT6B*, D+5)). In the remaining participant (participant 32 (*PPP1R12A*, A-21)), no disruption to splicing was observed (Table 1, Supplementary Figure 3). For two additional participants where the candidate variant fell in a canonical splice site (participants 83 (*TAOK1*, A-2) & 94 (*PHIP*, A-2)), a new diagnosis was reached without the need for functional work based on ACMG criteria with a PVS1 classification for these variants (Table 1, Supplementary Figure 3).

**Table 1.**
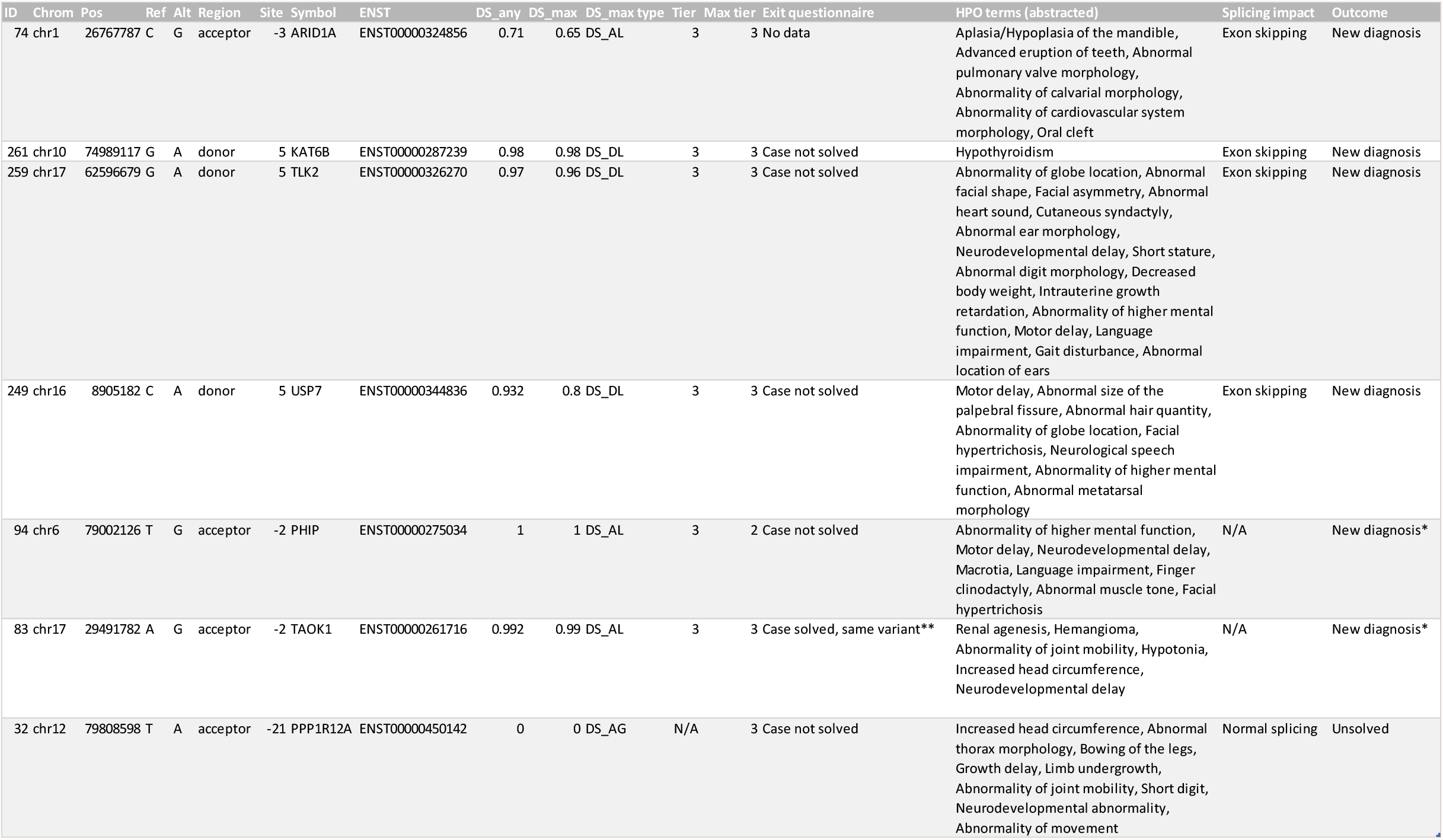
Diagnostic outcomes for seven individuals after clinical and functional characterisation of the splicing variant. Five individuals underwent RNA studies, of which four received a new diagnosis. In two additional individuals, a diagnosis was reached without the need for RNA studies. In total, a new diagnosis was confirmed for six individuals. Note that the given HPO terms are “abstracted “ (see Methods) to protect confidentiality. * In participants 83 and 94, a new diagnosis was reached without the need for functional evaluation. ** This exit questionnaire outcome was updated after the participant was identified by this study.

In summary, we demonstrate a functional splicing defect in four out of five participants recruited to our study, and we have identified a new molecular diagnosis for six individuals to date.

## Discussion

We examined WGS data from 38,688 individuals in the Rare Disease arm of the 100KGP to evaluate the contribution of splicing variants to rare genetic diseases. Using a population-based metric of constraint, MAPS, we showed that certain near-splice and (for the first time) branchpoint positions are under strong purifying selection. We identified 258 *de novo* near-splice and branchpoint variants in known disease genes in these families. We identified 35 likely diagnostic variants which had previously been missed through the 100KGP, and we have confirmed a new molecular diagnosis for six participants to date. Overall, we demonstrate the clinical value of examining both canonical and non-canonical splicing variants in unsolved rare diseases.

### Non-canonical splicing positions harbour deleterious splicing variants

We used three orthogonal approaches to estimate the deleteriousness of near-splice and branchpoint variants: between-species conservation, within-species constraint, and splicing pathogenicity prediction. The PhyloP, MAPS, and SpliceAI scores at splicing positions consistently highlight those non-canonical splicing positions (especially D0 and D+5) which are likely to harbour damaging variants. Indeed, three out of three D+5 variants in which we performed RNA studies caused exon skipping. Importantly, although we use a cohort of unaffected parents as a proxy for a normal population, the MAPS data we present is highly concordant with the strong signals of negative selection at which have been previously described in the ExAC and DDD datasets^8^.

Extending this analysis to splicing branchpoints, we find strong signals of negative selection at a subset of branchpoint positions. These results are consistent with other measures of constraint previously described at bovine and human branchpoints^35^. We also identified candidate diagnostic variants at these positions, and we are awaiting RNA samples to functionally characterise these variants. The disruption of splicing branchpoints may therefore make an important contribution to rare disease^10,11^, and a systematic analysis of *de novo* variation at branchpoints is an exciting future research opportunity.

The ACMG variant interpretation guidelines give special status to CSS variants as “very strong “ diagnostic candidates in disorders where LoF is a known disease mechanism^6^. This remains the case in more detailed guidance for the interpretation of LoF variants which has recently been introduced^36^. However, the deleteriousness of splicing variants is not binary, but on a continuum, and can be quantitively compared to other variant classes. Previous estimates suggest that 46% of non-canonical near-splice DNMs in dominant rare disease genes may be pathogenic, rising to 71% for pyrimidine to purine transversions in the polypyrimidine tract, and 90% for D+5 variants^8^. The deleteriousness of individual variants is contingent on many factors, such as local sequence context, the alternate nucleotide, exon frame, exon length, and intron length^9^. For this reason, the systematic classification of near-splice variants remains challenging, and clinical interpretation of these variants is still dependent on expert phenotype matching and functional validation of candidate variants.

The functional characterisation of splicing variants can be challenging and requires adequate amounts of good quality RNA. Our study is limited by the use of blood as a proxy for the most clinically relevant tissue, although we affirm the utility of blood RNA analysis by identifying splicing defects in four out of five samples tested. Whereas RT-PCR is a bespoke and low-throughput approach, going forward, RNA-sequencing (RNA-seq) offers an unbiased and high-throughput alternative to simultaneously detect and functionally characterise splicing variants. A whole-transcriptome RNA-seq pilot study has recently been proposed for 100KGP, and use of RNA-seq in routine clinical practice could offer a much-needed means to systematically and objectively interpret splicing variants^37^.

### New rare disease diagnoses

We identified 258 *de novo* SNVs overlapping near-splice or branchpoint regions of known monoallelic “loss of function “ rare disease genes in 255 individuals. Of these, at least 84 were already considered to be diagnostic through 100KGP, and we identified an additional 35 variants which are likely to be diagnostic given the available molecular, phenotypic, and *in silico* data. We confirmed a new molecular diagnosis for six participants, including four participants for whom RNA studies were performed.

Surprisingly, several strong diagnostic candidates were apparently overlooked in the standard variant interpretation pipeline, including at least nine CSS variants and four D+5 variants, all in known rare disease genes. Of ten *de novo* D+5 variants, none were previously labelled as pathogenic, despite their high prior probability of being diagnostic in this context^8^.

Clearly, many new diagnoses remain to be found. A recent analysis of 100KGP data in the context of craniosynostosis found that expert-led review more than doubled diagnostic yields compared to the standard pipeline^3^. An important factor is that the “virtual panels “ applied to variant calls are outdated and do not include recently discovered disease genes. Our phenotype-matching work suggests that the clinical impact of near-splice variants has been under-ascertained in this cohort, and we are continuing to recruit participants for functional assessment of these variants.

One obstacle to increasing the number of researcher-identified diagnoses in this context is the difficulty of recontacting de-identified participants and clinicians through secure research environments. The confidentiality of all participants in research is rightly a priority, and new pathways must be developed to streamline the clinical-research interface in medical genomics.

## Conclusion

In conclusion, the disruption of splicing is an important cause of rare disease among 100KGP participants, but the contribution of non-canonical variants is still under-recognised. Splicing branchpoints are another non-canonical and non-coding source of damaging splicing variants which are amenable to systematic analysis in WGS data. The improved interpretation of splicing variants is an area of great promise to genomic medicine and, above all, to individuals with rare diseases and their families.

## Data Availability

All code is available online at https://github.com/alexblakes/100KGP_splicing.
The data used in this study are available to registered users within a protected research environment at https://www.genomicsengland.co.uk/about-gecip/for-gecip-members/data-and-data-access/.

https://github.com/alexblakes/100KGP_splicing

## Supplemental data

Supplementary data include three figures and two tables.

## Acknowledgements

The authors thank all participants and families involved in this research. We thank all clinicians and contributors who helped to assess potential splicing diagnoses, including: Ellen Thomas, Jessica Radley, Rebecca Igbokwe, Suresh Vijay, Deirdre Cilliers, Evan Reid, Mick Parker, David Hunt, Rachel Keen, Ed Blair, Helen Firth, Peggy O ‘Driscoll, Chiara Marini Bettol, Monish Suri, John Barton, Angela Barnicoat, Sahar Mansour, Melody Redman, Kate Barr, Debbie Fuller, Meena Balasubramanian, Julia Rankin, Sian Ellard, Olga Tsoulaki, and Emma Kivuva.

The Baralle lab is supported by NIHR Research Professorship awarded to D.B. (RP-2016-07-011). Functional work was additionally supported by a Wessex Medical Research Innovation Grant awarded to J.L. NW is currently supported by a Sir Henry Dale Fellowship jointly funded by the Wellcome Trust and the Royal Society (Grant Number 220134/Z/20/Z) and funding from the Rosetrees Trust. AB was supported by funding from Health Education England. We acknowledge the NIHR Clinical Research Network (CRN) in recruiting participants and the Musketeers Memorandum, as well as support from the NIHR UK Rare Genetic Disease Consortium. We thank Patrick J. Short for providing a curated set of de novo variants used in earlier iterations of these analyses.

This research was made possible through access to the data and findings generated by the 100,000 Genomes Project. The 100,000 Genomes Project is managed by Genomics England Limited (a wholly owned company of the Department of Health and Social Care). The 100,000 Genomes Project is funded by the National Institute for Health Research and NHS England. The Wellcome Trust, Cancer Research UK and the Medical Research Council have also funded research infrastructure. The 100,000 Genomes Project uses data provided by participants and collected by the National Health Service as part of their care and support.

## Data and code availability

All code is available online at https://github.com/alexblakes/100KGP_splicing.

The data used in this study are available to registered users within a protected research environment at https://www.genomicsengland.co.uk/about-gecip/for-gecip-members/data-and-data-access/.

## Supplementary Figures

Supplementary Figure 1

Tiering data for splicing DNMs in known monoallelic loss-of-function rare disease genes. Each point represents a DNM in a rare disease proband. Points are coloured by the “tier “ of that variant in the GEL annotation pipeline (see Methods).

Supplementary Figure 2

SpliceAI scores of splicing DNMs in known monoallelic loss-of-function rare disease genes. Each point represents a DNM in a rare disease proband. Points are coloured by SpliceAI score; grey points indicate that no SpliceAI annotation is available.

Supplementary Figure 3

Functional outcomes for participant samples which were characterised by RT-PCR. Each page illustrates the following: a schematic of variant position relative to the exon/intron junction, a schematic of the splicing consequence of each variant, gel electrophoresis of amplified RT-PCR products for participant and control samples, Sanger sequencing trace for proband-specific bands.

## Supplementary tables

Supplementary Table 1

Molecular details and clinical outcome data for the 258 prioritised DNMs. Rows in bold indicate those variants identified as likely to be diagnostic, which are given in Supplementary Table 2 (“Main “ DNM set). * This exit questionnaire outcome was updated after the participant was identified by this study.

Supplementary Table 2

Molecular details and phenotypic data for 35 likely diagnostic variants in “unsolved “ probands. Each variant was judged to be at least a plausible diagnostic fit given the phenotype information available. Note that the given HPO terms are “abstracted “ (see Methods) to prevent participant re-identification. Also note that of the 35 diagnostic candidates, 8 were identified in a preliminary set of DNMs used for exploratory analysis at the inception of this project, and are not among the 258 prioritised DNMs (Supplementary Table 1) described in more detail above (see Methods). Participants in bold are those shown in Table 1. * This exit questionnaire outcome was updated after the participant was identified by this study.

