## Supplementary figures and images for "A systematic analysis of splicing variants identifies new diagnoses in the 100,000 Genomes Project"

### Supplementary Figure 1

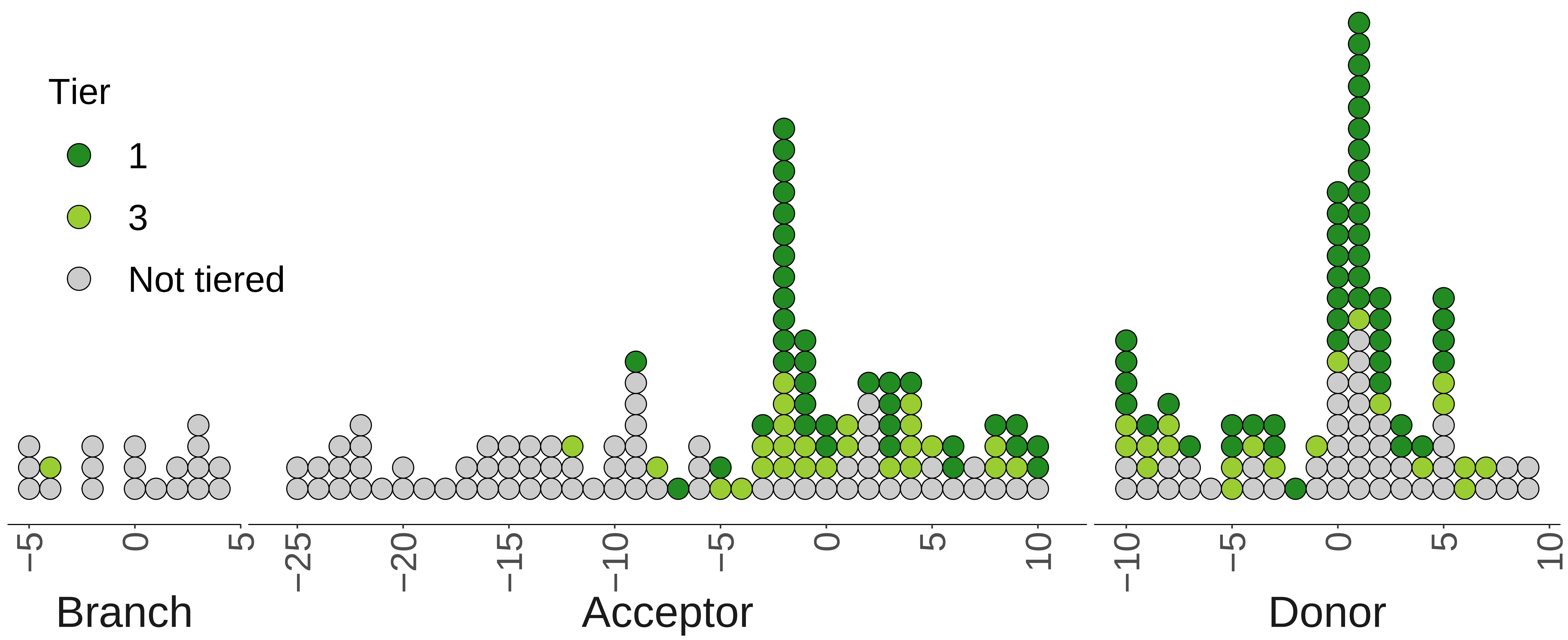

### Supplementary Figure 2

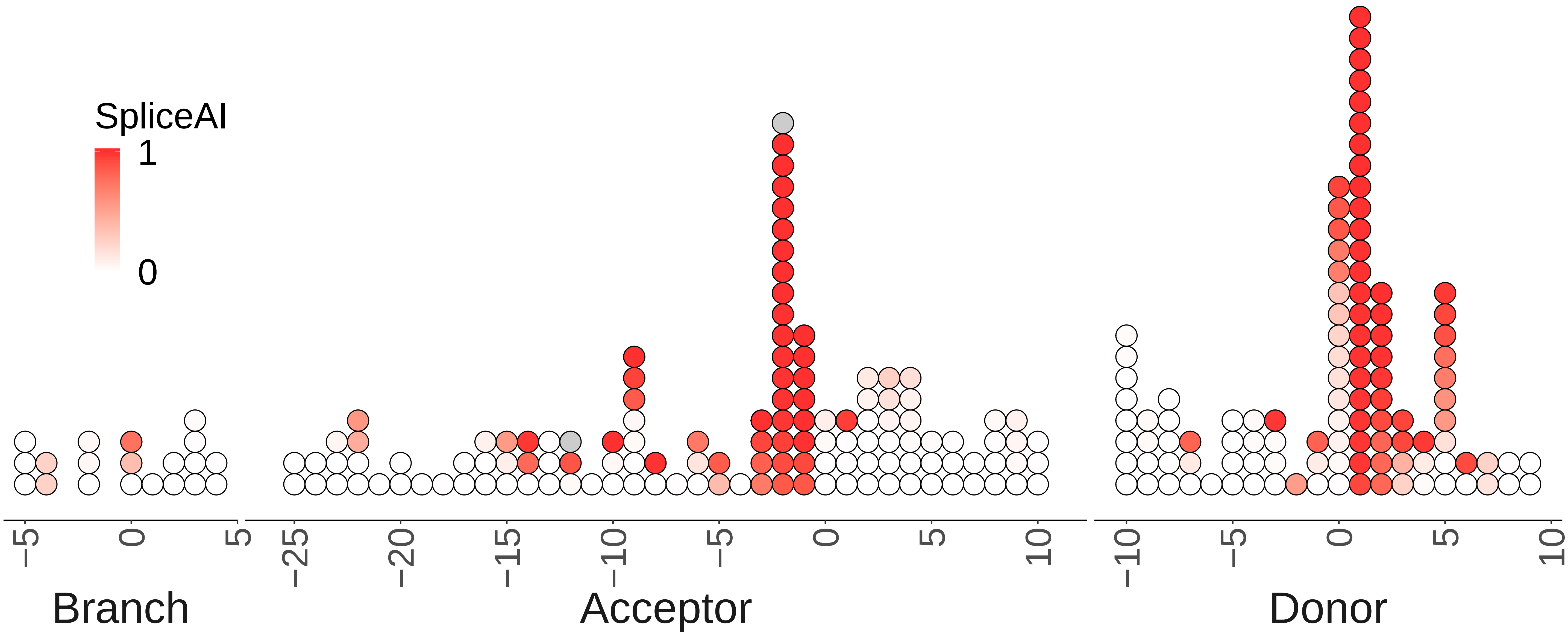
