## Supplementary Figure 3 for "A systematic analysis of splicing variants identifies new diagnoses in the 100,000 Genomes Project"

**Supplemental Figure X: Outcomes of RT-PCR functional validations of splicing variants**

| Variant, gene | chr10:74989117 G>A, *KAT6B* |
|---|---|
| Sequence schematic | 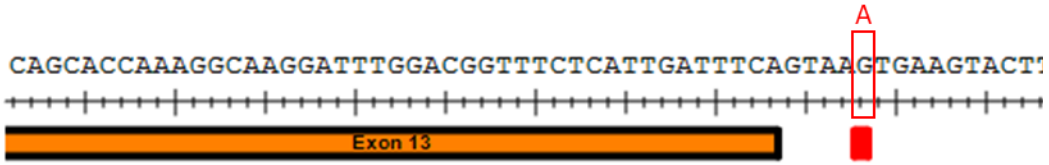<br> |
| Splicing schematic | 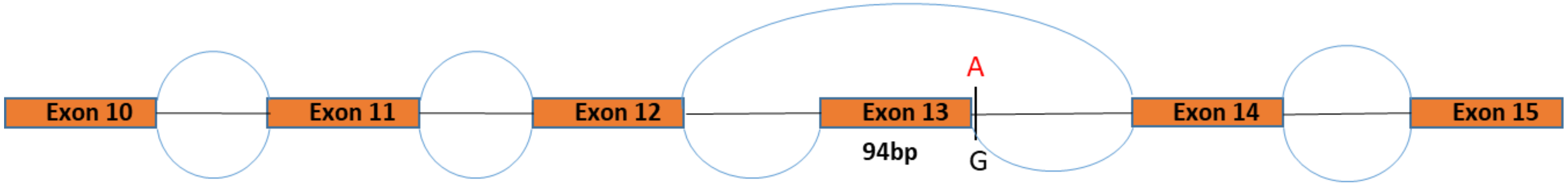<br> |
| Gel electrophoresis | 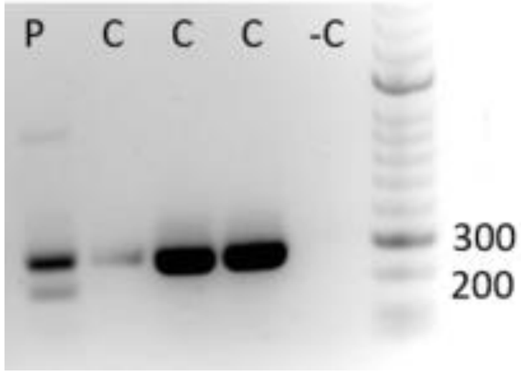<br><br>P=Proband, C=Control, -C=DNA free control |
| Sanger sequencing proband specific band | 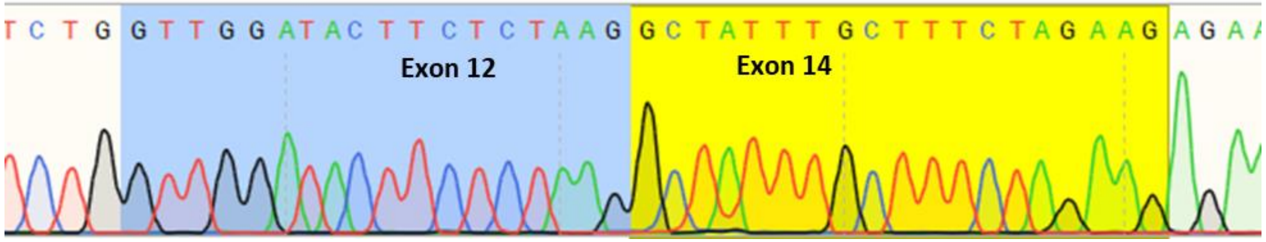<br><br>Exon 13 skipped |

| Variant, gene | chr17:62596679 G>A, *TLK2* |
|---|---|
| Sequence schematic | 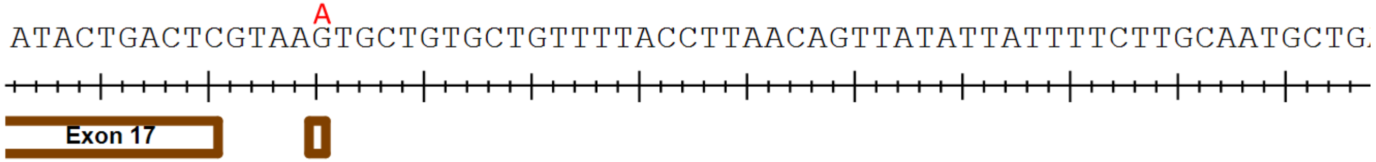  |
| Splicing schematic | 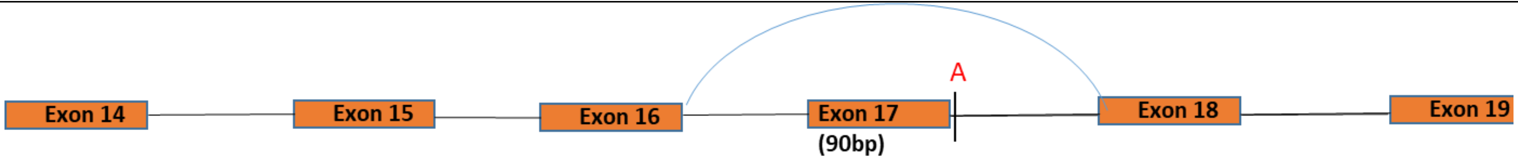  |
| Gel electrophoresis | 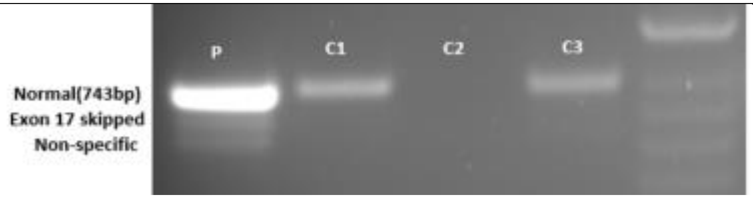 <br>P=Proband, C=Control |
| Sanger sequencing proband specific band | 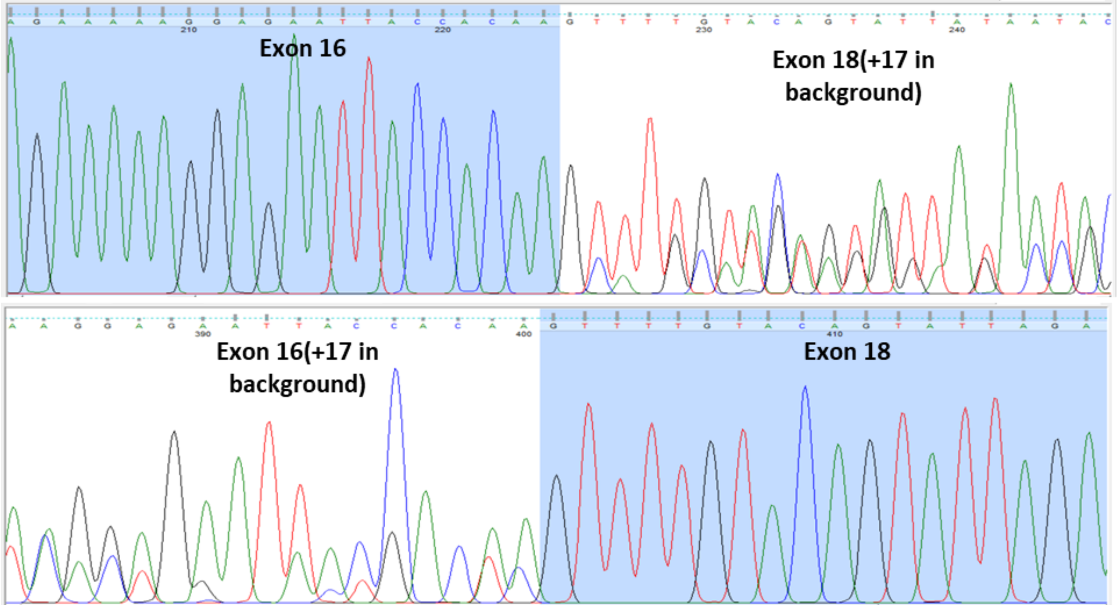 <br>Exon 17 skipped |

| | |
|---|---|
| Variant, gene | chr1:26767787 C>G, *ARID1A* |
| Sequence schematic | 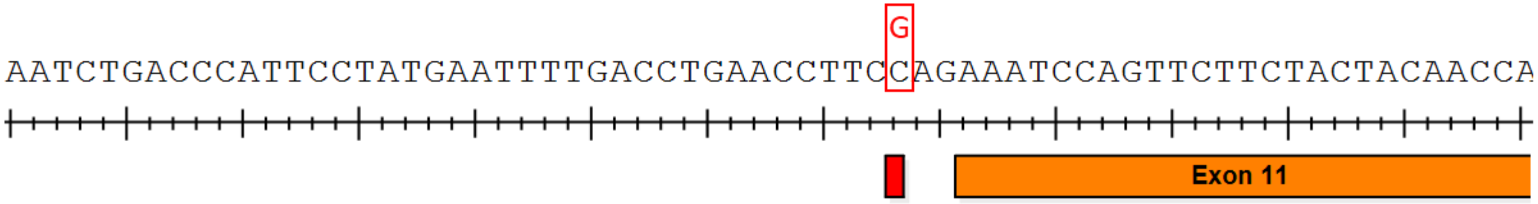  |
| Splicing schematic | 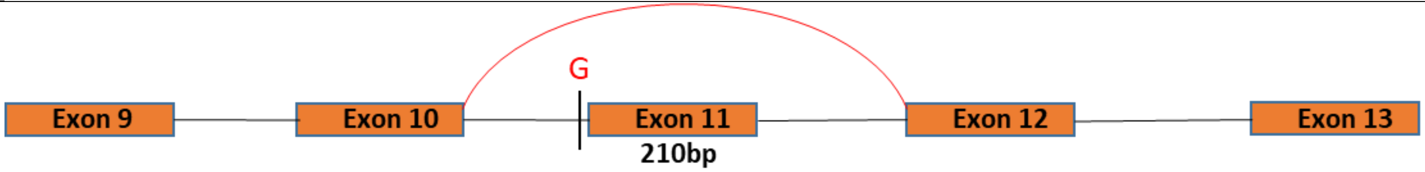  |
| Gel electrophoresis | 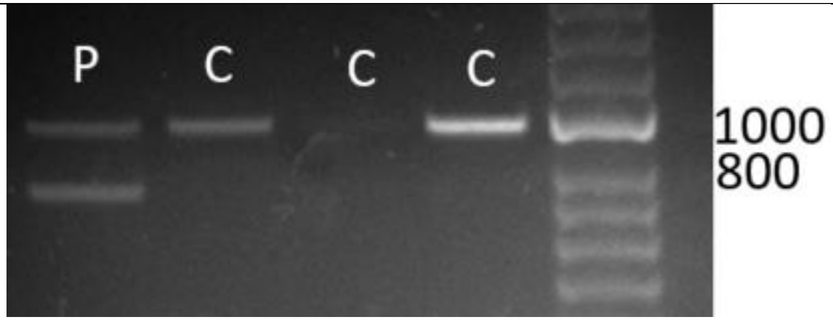  P=proband, C=Control |
| Sanger sequencing proband specific band | 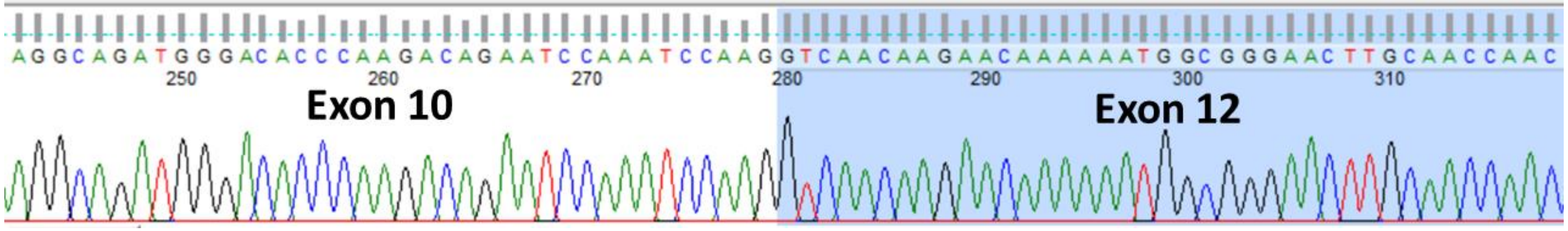  Exon 11 skipped |

| Variant, gene | chr16:8905182 C>A, *USP7* |
|---|---|
| Sequence schematic | 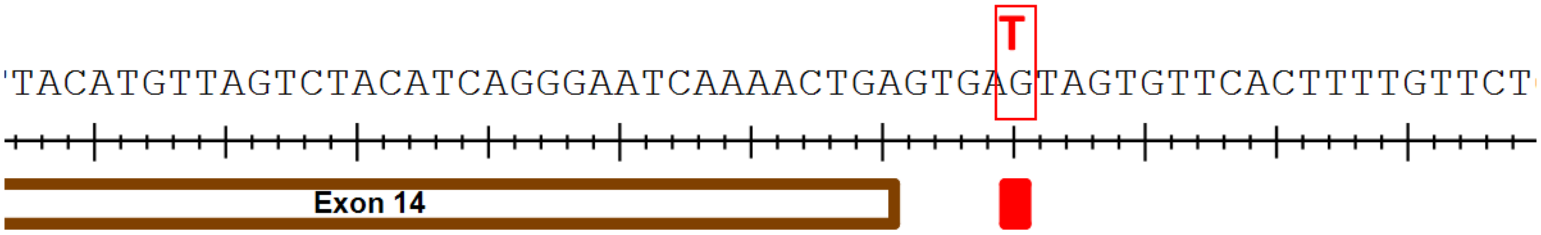 |
| Splicing schematic | 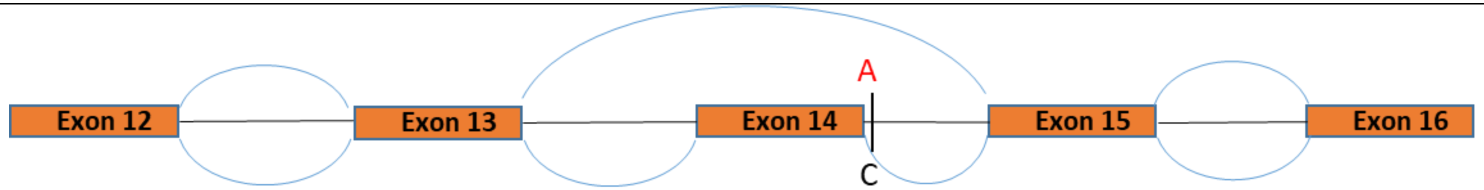 |
| Gel electrophoresis | 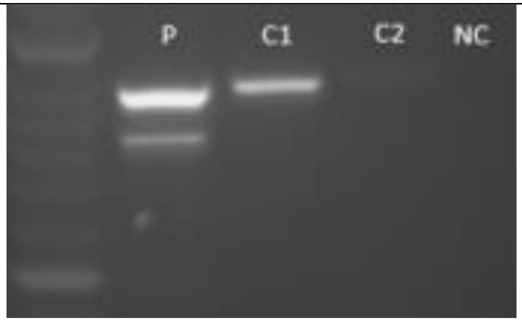<br>P=proband, C=Control, NC = no template control |
| Sanger sequencing proband specific band | 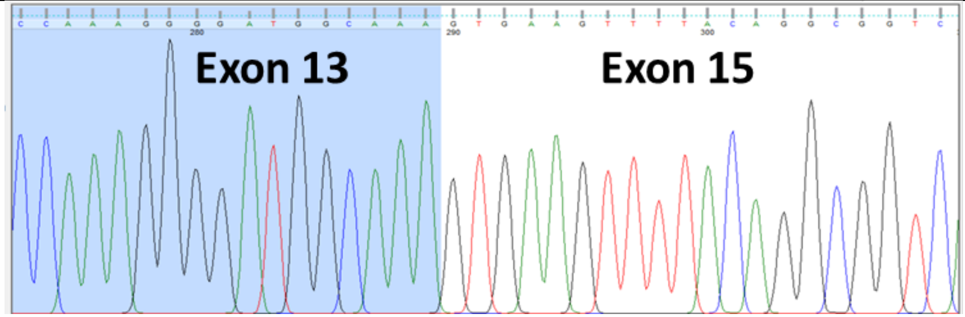<br>Exon 14 skipped |

<table>
<tr><td>Variant, gene</td><td>chr12:79808598 T>A, *PPP1R12A*</td></tr>
<tr><td>Sequence schematic</td><td>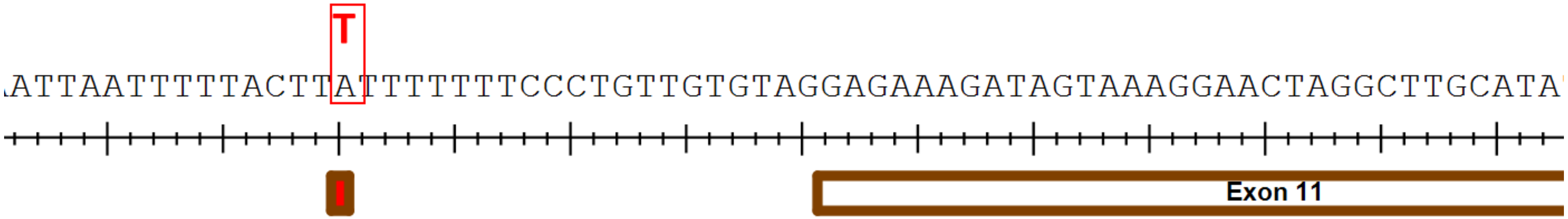
</td></tr>
<tr><td>Splicing schematic</td><td>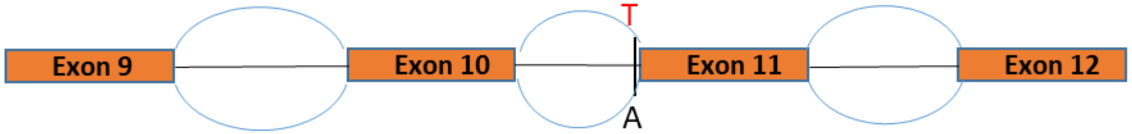
</td></tr>
<tr><td>Gel electrophoresis</td><td>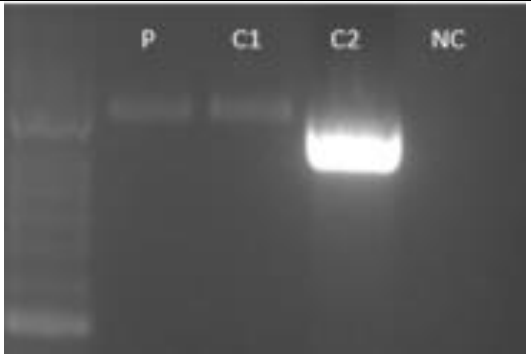

P=proband, C=Control, NC = no template control</td></tr>
<tr><td>Sanger sequencing</td><td>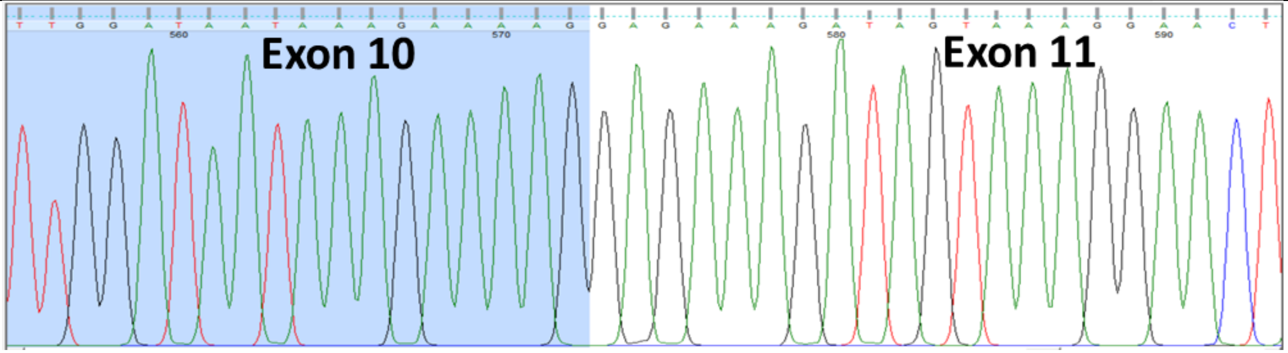

Normal splicing</td></tr>
</table>
